# Timing of Breakthrough Infection Risk After Vaccination Against SARS-CoV-2

**DOI:** 10.1101/2022.01.04.22268773

**Authors:** David N. Fisman, Nelson Lee, Ashleigh R. Tuite

## Abstract

**Background:** Provision of safe and effective vaccines has been a remarkable public health achievement during the SARS-CoV-2 pandemic. The effectiveness and durability of protection of the first two doses of SARS-CoV-2 vaccines is an important area for study, as are questions related to optimal dose combinations and dosing intervals.

**Methods:** We performed a case-cohort study to generate real-world evidence on efficacy of first and second dose of SARS-CoV-2 vaccines, using a population-based case line list and vaccination database for the province of Ontario, Canada between December 2020 and October 2021. Risk of infection after vaccination was evaluated in all laboratory-confirmed vaccinated SARS-CoV-2 cases, and a 2% sample of vaccinated controls, evaluated using survival analytic methods, including construction of Cox proportional hazards models. Vaccination status was treated as a time-varying covariate.

**Results:** First and second doses of SARS-CoV-2 vaccine markedly reduced risk of infection (first dose efficacy 68%, 95% CI 67% to 69%; second dose efficacy 88%, 95% CI 87 to 88%). In multivariable models, extended dosing intervals were associated with lowest risk of breakthrough infection (HR for redosing 0.64 (95% CI 0.61 to 0.67) at 6-8 weeks). Heterologous vaccine schedules that mixed viral vector vaccine first doses with mRNA second doses were significantly more effective than mRNA only vaccines. Risk of infection largely vanished during the time period 4-6 months after the second vaccine dose, but rose markedly thereafter.

**Interpretation:** A case-cohort design provided an efficient means to identify strong protective effects associated with SARS-CoV-2 vaccination, particularly after the second dose of vaccine. However, this effect appeared to wane once more than 6 months had elapsed since vaccination. Heterologous vaccination and extended dosing intervals improved the durability of immune response.

## Introduction

The rapid introduction of highly effective vaccines against SARS-CoV-2 represents a remarkable scientific achievement amidst the current global pandemic (1). It has been estimated that these vaccines have averted hundreds of thousands of deaths (2, 3). While mRNA vaccines represent the first application of a novel and highly immunogenic vaccine construct, viral vector vaccines have also been extremely effective in preventing severe illness and death (4). The introduction of novel vaccines against a virus that has been recognized for only two years (5), means that there is of necessity a great deal of uncertainty related to their application. The relative protection afforded by vaccines against emerging variants of concern (6), vaccine safety (7, 8), the immunogenicity of mixed vaccine schedules (9), optimal spacing between administration of doses (10), the number of doses required for a full primary series (11), and the necessity of future boosting (12), are all the subject of active study.

The Canadian province of Ontario represents a large (population 14.6 million) and diverse jurisdiction, with high levels of 2-dose SARS-CoV-2 vaccine coverage (approximately 77% as of December 2021) (13, 14). The province has made use of several different vaccine formulations, predominantly BNT162b2 (BNT/Pfizer, “Cominrty”), mRNA-1273 (Moderna, “SpikeVax”), and ChAdOX1-S (AstraZeneca, “Covishield”). Ontario’s vaccination program has prioritized risk groups over time. For example, the program initially focussed on vaccinating residents of long-term care facilities and health-care workers (15). Once higher risk groups were vaccinated, eligibility was expanded using a descending age-based approach. The province also consciously chose to offer extended dosing intervals to lower risk populations, due to supply constraints and the potential advantages of rapid, maximal coverage with first doses of vaccine (16). Pediatric vaccination programs for those aged 5 to 11 years were introduced in in late November 2021 (17).

Beginning in autumn 2021, third doses have been offered to a growing list of individuals identified as being at elevated risk due to age, medical comorbidity or congregate living residence. Although there was some evidence suggesting that the use of prolonged dosing intervals in the primary series had resulted in durable immunity that made third doses unnecessary (18), the province expanded access to third vaccine to all aged 50 and over December 2021 (19). In late December, the rapid spread of the Omicron variant of concern in the province resulted in an expansion of third dose eligibility to all aged 18 and over who received their second dose at least 3 months previously.

A large, highly-vaccinated population like Ontario’s, which has received a several different vaccination regimens, with variable dosing intervals, provides an opportunity to generate real-world evidence on efficacy of vaccination against breakthrough infections. The province’s vaccination database and COVID-19 case line provide detailed information on vaccine administration as well as individual-level covariates that can be used to adjust for confounding by factors associated with both infection risk and vaccination priority. Our objective was to use these data to generate estimates of 1- and 2-dose vaccination protection over time, against breakthrough SARS-CoV-2 infection; we also performed exploratory analyses to generate hypotheses related to optimal dosing intervals and the relative efficacy of different vaccine combinations.

## Methods

### Data Sources

We created a cohort of individuals who received vaccination against SARS-CoV-2 in Ontario, without record of infection prior to the date of receipt of first vaccine dose. We included all reported cases of infection meeting these criteria, as well as a 2% random sample of vaccinated individuals with no record of infection. Cases were identified in the Province’s Case and Contact Management (CCM) database as described elsewhere (20, 21); we included only cases with a unique “pseudo-health card number”, which permitted linkage with the provincial vaccination database. Vaccination information on cases and controls was extracted from the Province’s COVaxON dataset (21), which includes dosage dates, vaccines used, basic demographic information, and fields on indication for vaccination, including residence in a congregate setting (including residence in a nursing home, retirement community, or other congregate setting), priority health condition, and healthcare worker status. As with cases, the pool of potential controls was limited to individuals with a pseudo-health card number. A flow diagram outlining creation of the cohort is presented in **Figure 1**.

**Figure 1.**
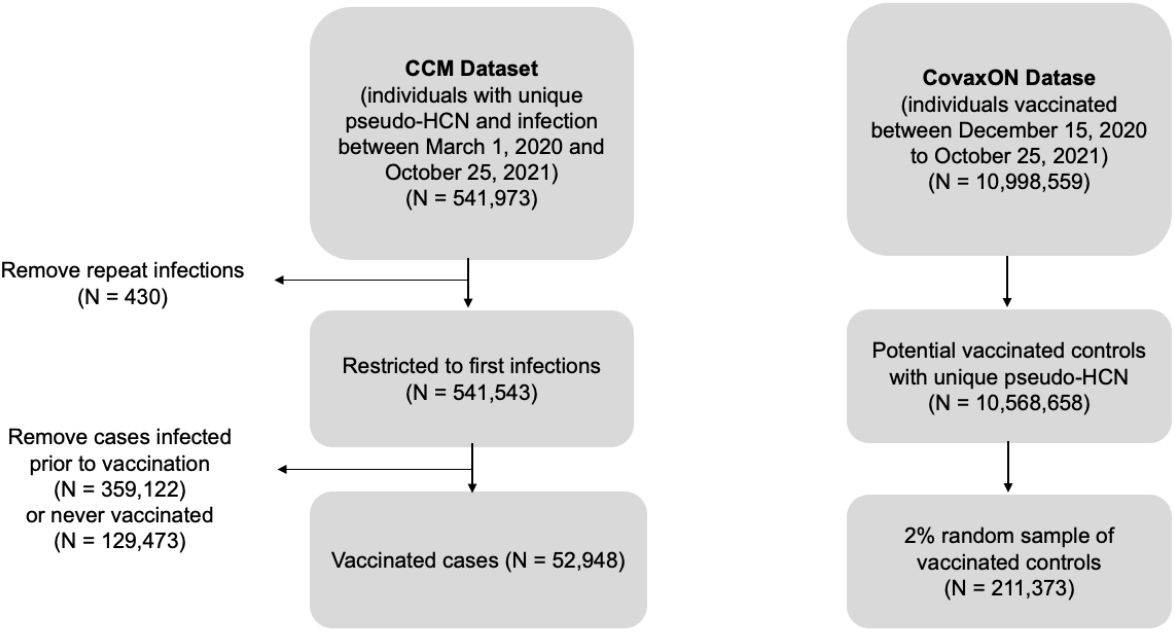
Flow Diagram for Creation of Case-Cohort Sample.

We considered individuals to have been vaccinated with a first dose of vaccine during time at risk 14 or more days after the date of the first vaccine dose; individuals were considered vaccinated with two doses of vaccine during time at risk 14 or more days after second vaccine dose. While some researchers, including ourselves (21-23), have previously defined two dose-vaccinated time as commencing 7 or more days after second dose, we use the longer window for consistency with Canadian public health authorities (24).

### Analysis

We performed a case-cohort analysis using survival analytic methods. As person time data were available, we were able to estimate hazard ratios directly using Cox proportional hazards models, rather than approximate such ratios via risk set sampling. Controls were reweighted by a factor of 50 to account for 2% selection probability. We stratified within-person time at risk according to an individual’s vaccination status, which was treated as a time-varying covariate.

We created Cox proportional hazards models including only vaccination as an exposure (unvaccinated, single-dose vaccinated, or fully vaccinated), as well as adjusted models that adjusted for variables that we expected to confound observed vaccine effect. We evaluated the proportional hazards assumption both graphically using log-log plots, and statistically based on Schoenfeld residuals (25). As we found hazards of infection following vaccination to be non-proportional through inspection of log-log plots and testing of Schoenberg residuals (**Supplementary Figure**), we re-ran our Cox proportional hazards models with time at risk further stratified by 2-month blocks, within a given vaccination status.

Lastly, we performed restriction analyses in which we restricted cases individuals with infection due to a non-variant of concern viral strain, to infection with N501Y+ variants of concern (alpha, beta and gamma variants), and to infection with likely delta infection. Classification of likely variant status of the infecting strain was as described previously (21). We conducted all analyses in Stata version 15.1 using sts, stox and related commands. The study was conducted in accordance with the STROBE guidelines for observational research (26), and received ethics approval from the Research Ethics Board at the University of Toronto.

## Results

Our final cohort consisted of 52,948 cases and 211,373 controls. Cases and controls differed significantly according to age distribution, sex, vaccine combination received, documented priority health condition, residence in a congregate setting, and healthcare worker status, as well as interval between first and second doses (**Table 1**). Crude time to breakthrough infection by vaccination status is presented as a Kaplan-Meier curve (**Figure 2**); vaccination was associated with significant reduction in infection risk, with 2-dose vaccination reducing risk relative to single dose vaccination (P < 0.001 by log-rank test for all comparisons).

**Table 1:**
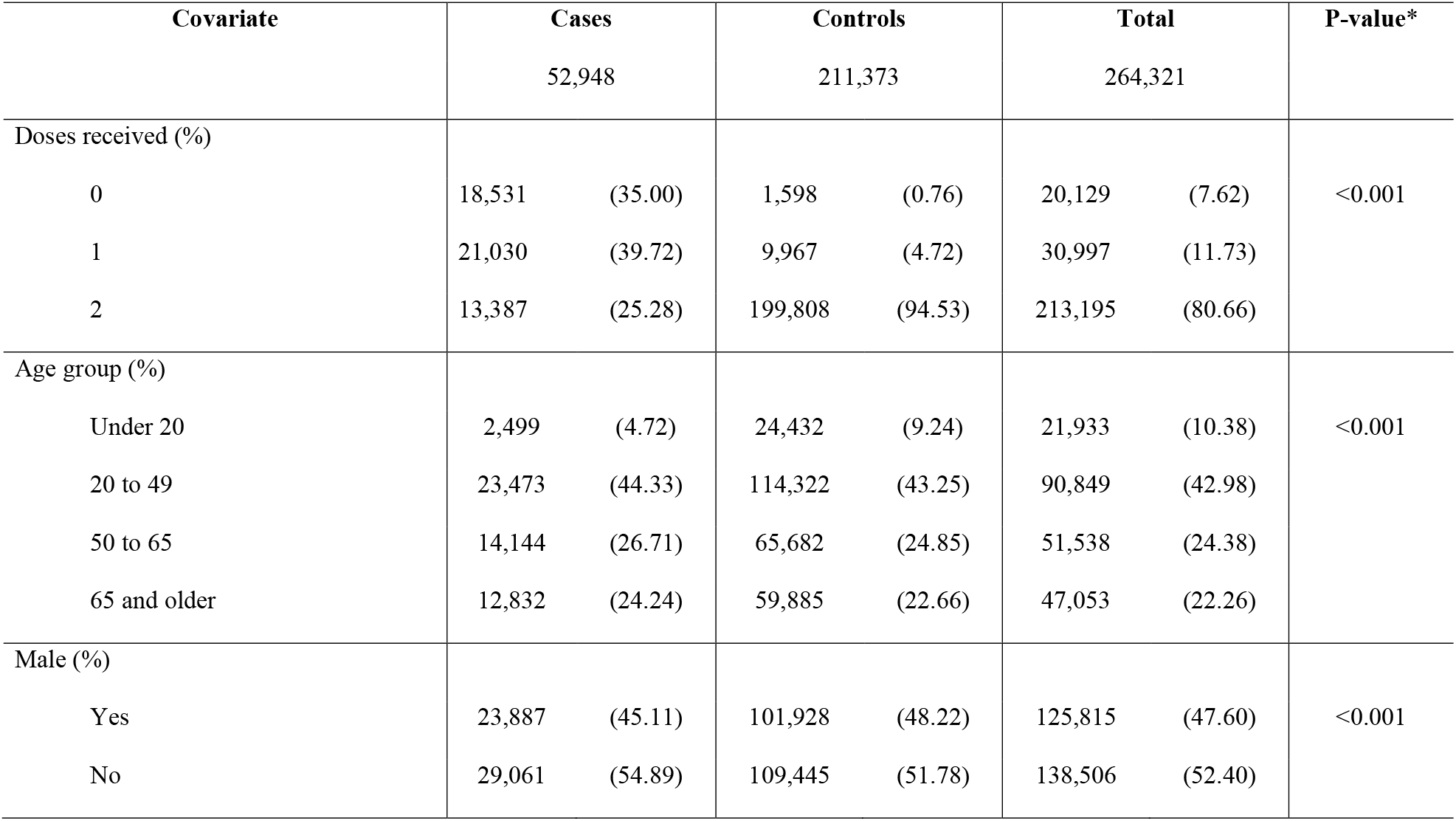
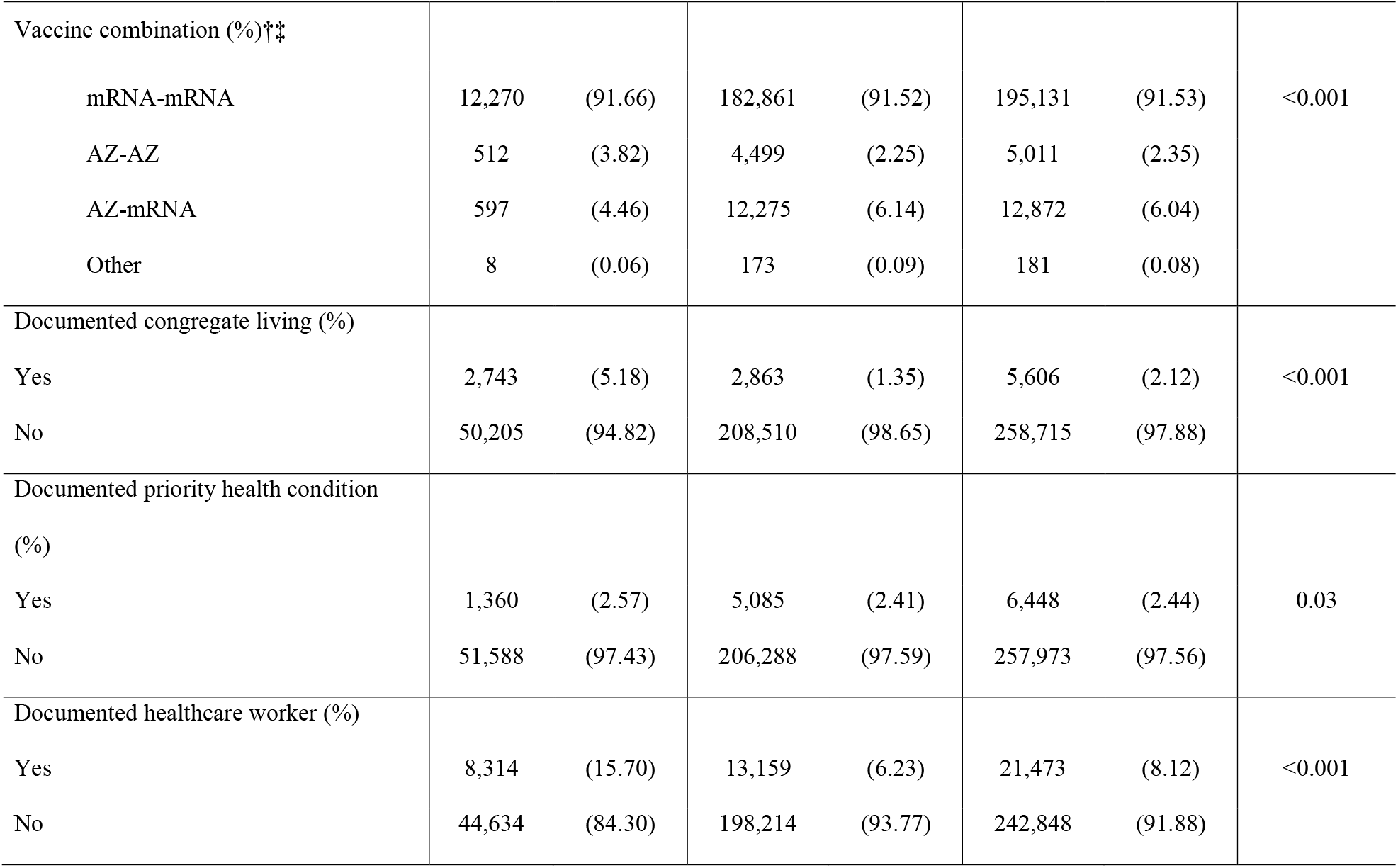
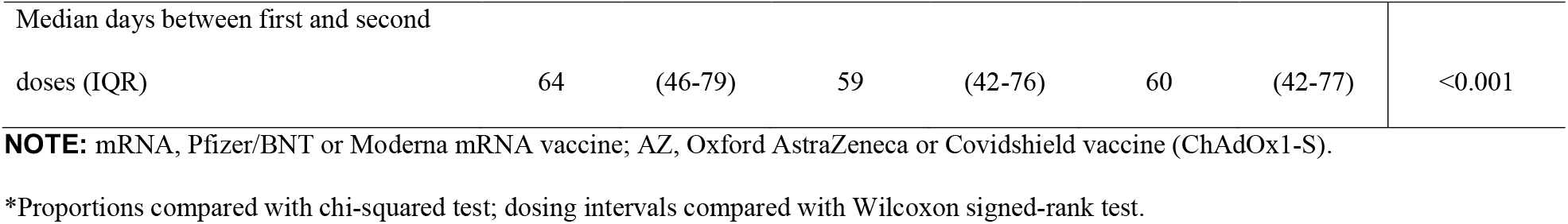
Characteristics of SARS-CoV-2-Vaccinated Cases and Controls, Ontario, Canada.

**Figure 2.**
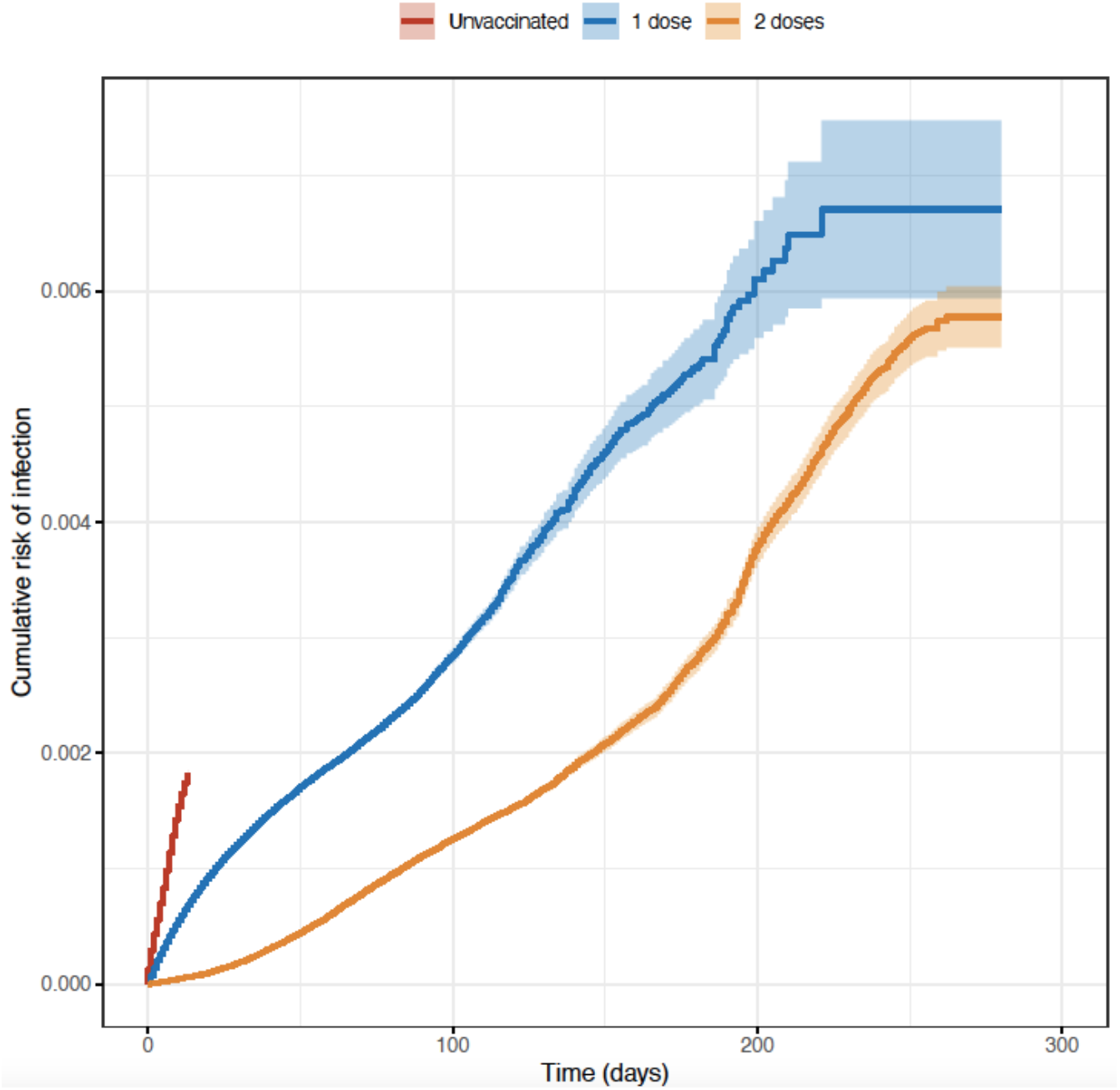
Time to Infection Without SARS-CoV-2 Vaccination, or After 1 or 2 Vaccine Doses. Vaccination status is treated as a time-varying covariate and defined by time since vaccination and cumulative doses, as described in the text. X-axis, person-time; Y-axis, cumulative incidence of infection. Shaded areas represent 95% confidence intervals.

We constructed a Cox proportional hazards model that included only vaccination status; the hazard ratio for breakthrough infection after a single dose of vaccine was 0.32 (95% CI 0.31 to 0.33); after a second dose of vaccine the hazard ratio declined to 0.12 (95% CI 0.12 to 0.13), corresponding to effectiveness of 68% and 88% against infection. We found little change in these after adjusting for potential confounders in multivariable models (**Table 2**). In multivariable analyses we identified a strong U-shaped relationship between dosing interval and risk of breakthrough infection, with risk lowest when vaccine doses were administered 6-8 weeks apart. Among individuals who received two doses of vaccine, increased risk of breakthrough infection was seen in those who received two doses of ChAdOx1-S, relative to two doses of mRNA vaccine. However, for individuals who received an initial dose of ChAdOx1-S, there was a significant reduction in breakthrough risk following a second dose of mRNA vaccine, in contrast to receipt of two doses of mRNA vaccine. Risk of breakthrough was highest in younger adults (aged 20-49), and was elevated in healthcare workers, those residing in congregate living settings, and in those identified as having priority medical conditions.

**Table 2.**
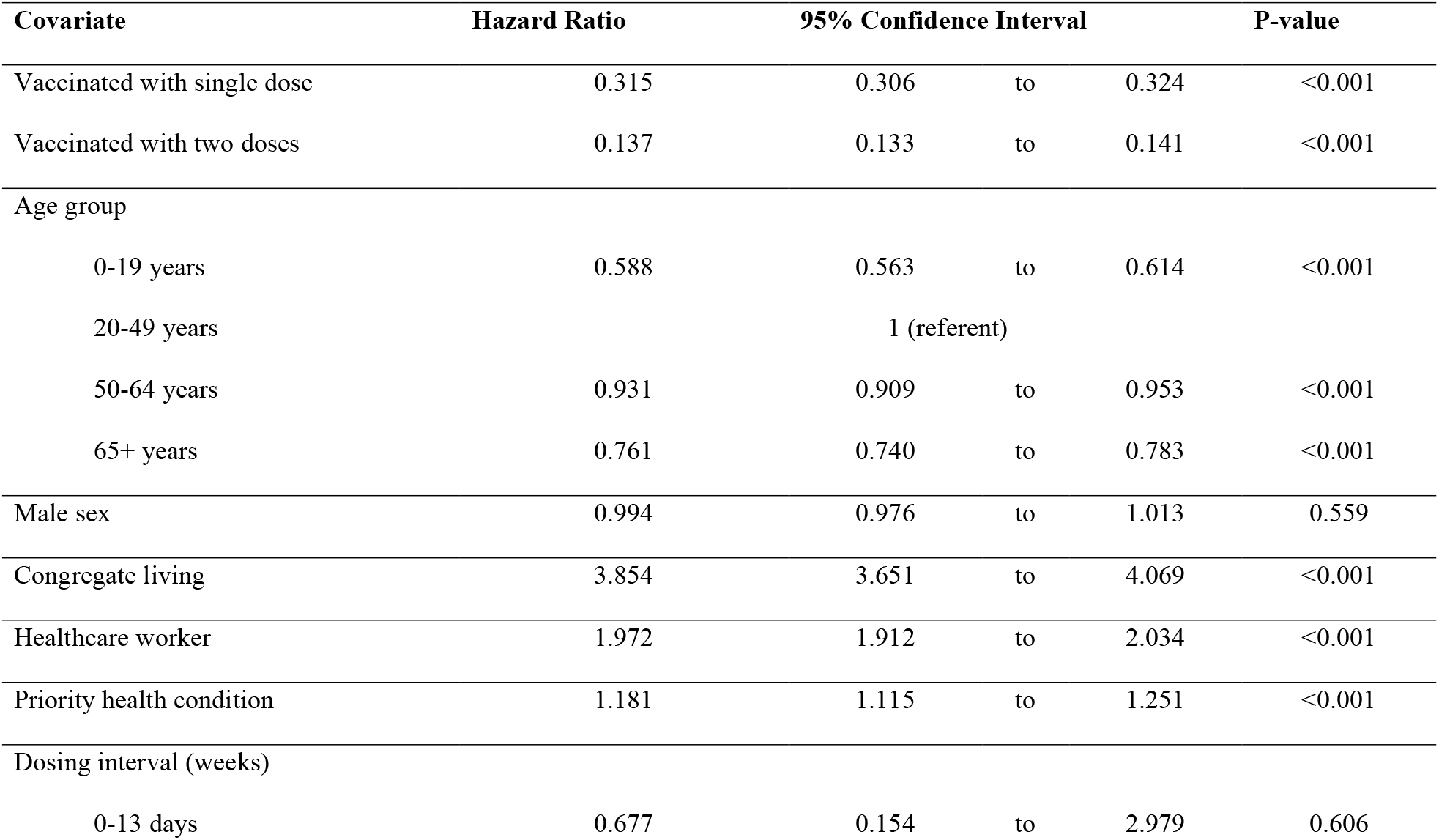
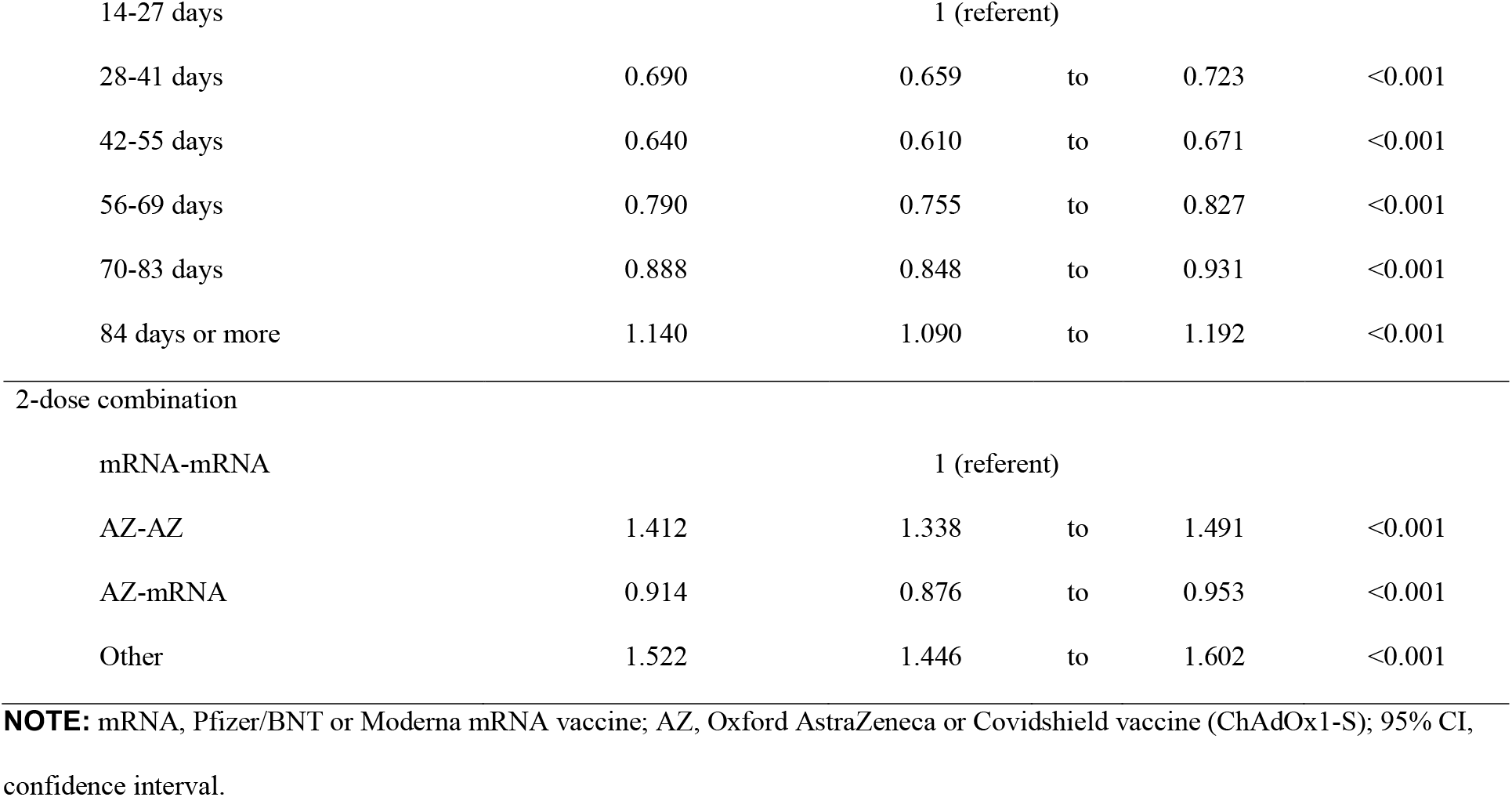
Results of Multivariable Cox Proportional Hazards Model.

| **Covariate** | **Hazard Ratio** | **95% Confidence Interval** |  |  | **P-value** |
| --- | --- | --- | --- | --- | --- |
| Vaccinated with single dose | 0.315 | 0.306 | to | 0.324 | <0.001 |
| Vaccinated with two doses | 0.137 | 0.133 | to | 0.141 | <0.001 |
| Age group |  |  |  |  |  |
| 0-19 years | 0.588 | 0.563 | to | 0.614 | <0.001 |
| 20-49 years |  | 1 (referent) |  |  |  |
| 50-64 years | 0.931 | 0.909 | to | 0.953 | <0.001 |
| 65+ years | 0.761 | 0.740 | to | 0.783 | <0.001 |
| Male sex | 0.994 | 0.976 | to | 1.013 | 0.559 |
| Congregate living | 3.854 | 3.651 | to | 4.069 | <0.001 |
| Healthcare worker | 1.972 | 1.912 | to | 2.034 | <0.001 |
| Priority health condition | 1.181 | 1.115 | to | 1.251 | <0.001 |
| Dosing interval (weeks) |  |  |  |  |  |
| 0-13 days | 0.677 | 0.154 | to | 2.979 | 0.606 |
| 14-27 days |  | 1 (referent) |  |  |  |
| 28-41 days | 0.690 | 0.659 | to | 0.723 | <0.001 |
| 42-55 days | 0.640 | 0.610 | to | 0.671 | <0.001 |
| 56-69 days | 0.790 | 0.755 | to | 0.827 | <0.001 |
| 70-83 days | 0.888 | 0.848 | to | 0.931 | <0.001 |
| 84 days or more | 1.140 | 1.090 | to | 1.192 | <0.001 |
| 2-dose combination |  |  |  |  |  |
| mRNA-mRNA |  | 1 (referent) |  |  |  |
| AZ-AZ | 1.412 | 1.338 | to | 1.491 | <0.001 |
| AZ-mRNA | 0.914 | 0.876 | to | 0.953 | <0.001 |
| Other | 1.522 | 1.446 | to | 1.602 | <0.001 |

**NOTE:** mRNA, Pfizer/BNT or Moderna mRNA vaccine; AZ, Oxford AstraZeneca or Covidshield vaccine (ChAdOx1-S); 95% CI, confidence interval.

Models that stratified time at risk both by vaccination status and 60-day time blocks since vaccination generated similar results to initial multivariable models, demonstrating high efficacy of first and second vaccine doses (**Supplementary Table**). The lowest hazard ratio for breakthrough infection was seen between four and 6 months after second vaccine dose but increased after 6 months of observation (**Figure 3**). In exploratory restriction analyses in which we included only cases with non-variant of concern infection, cases with N501Y+ infection (alpha, beta or gamma variants), or cases with delta variant infection, we found significant protection against breakthrough infection with both 1 and 2 doses of vaccine, against all variants. However, there was significant heterogeneity across variants (P for heterogeneity < 0.001 for both 1 and 2 doses), with highest efficacy after two doses seen against N501Y+ infection (96.9%, 95% CI 96.7% to 97.1%), and lower efficacy seen against delta variant (66.1%, 95% CI 62.5% to 69.4%) (**Table 3**).

**Table 3.**
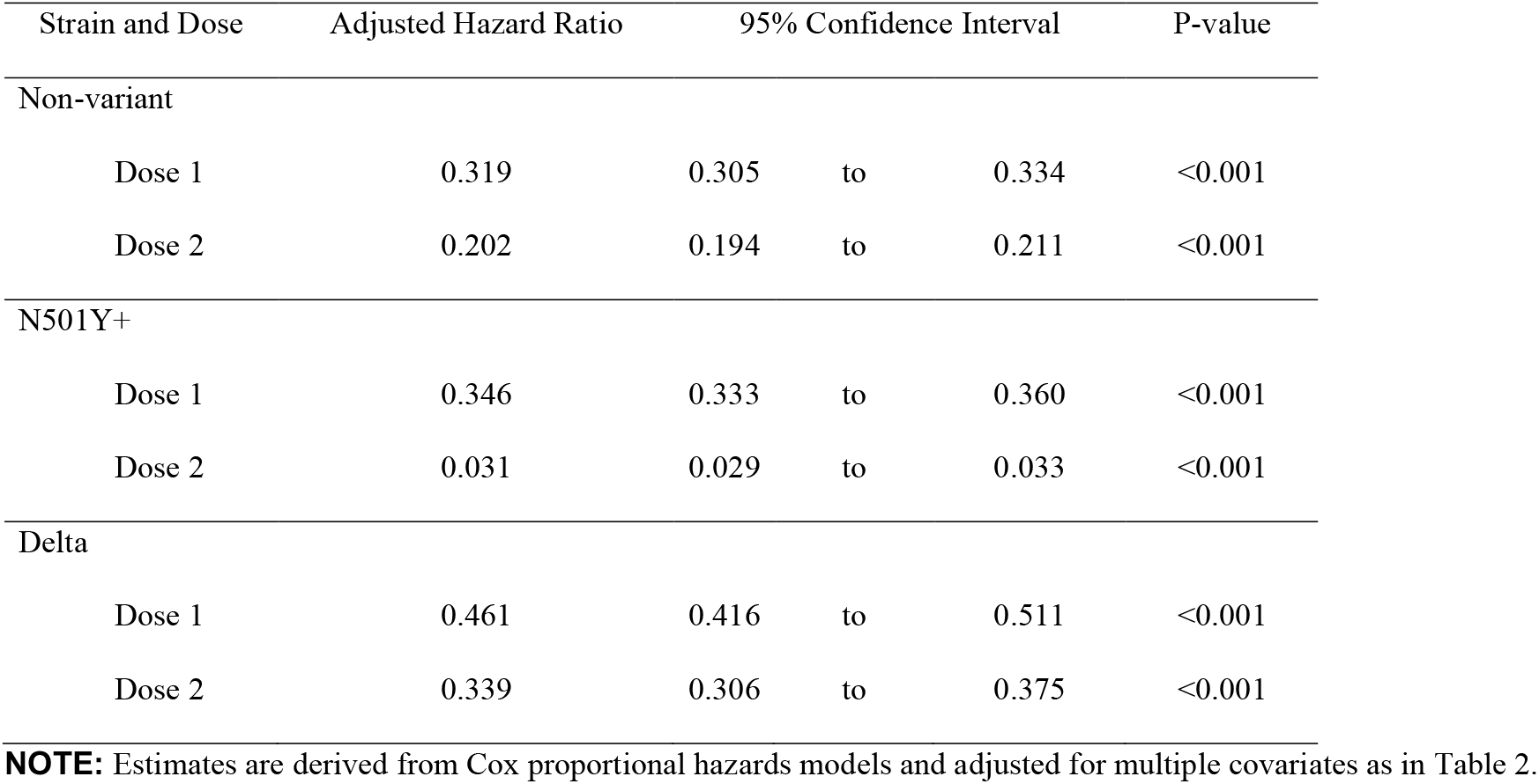
Strain- and Dose-Specific Efficacy of SARS-CoV-2 Vaccines in Restriction Analyses.

**Figure 3.**
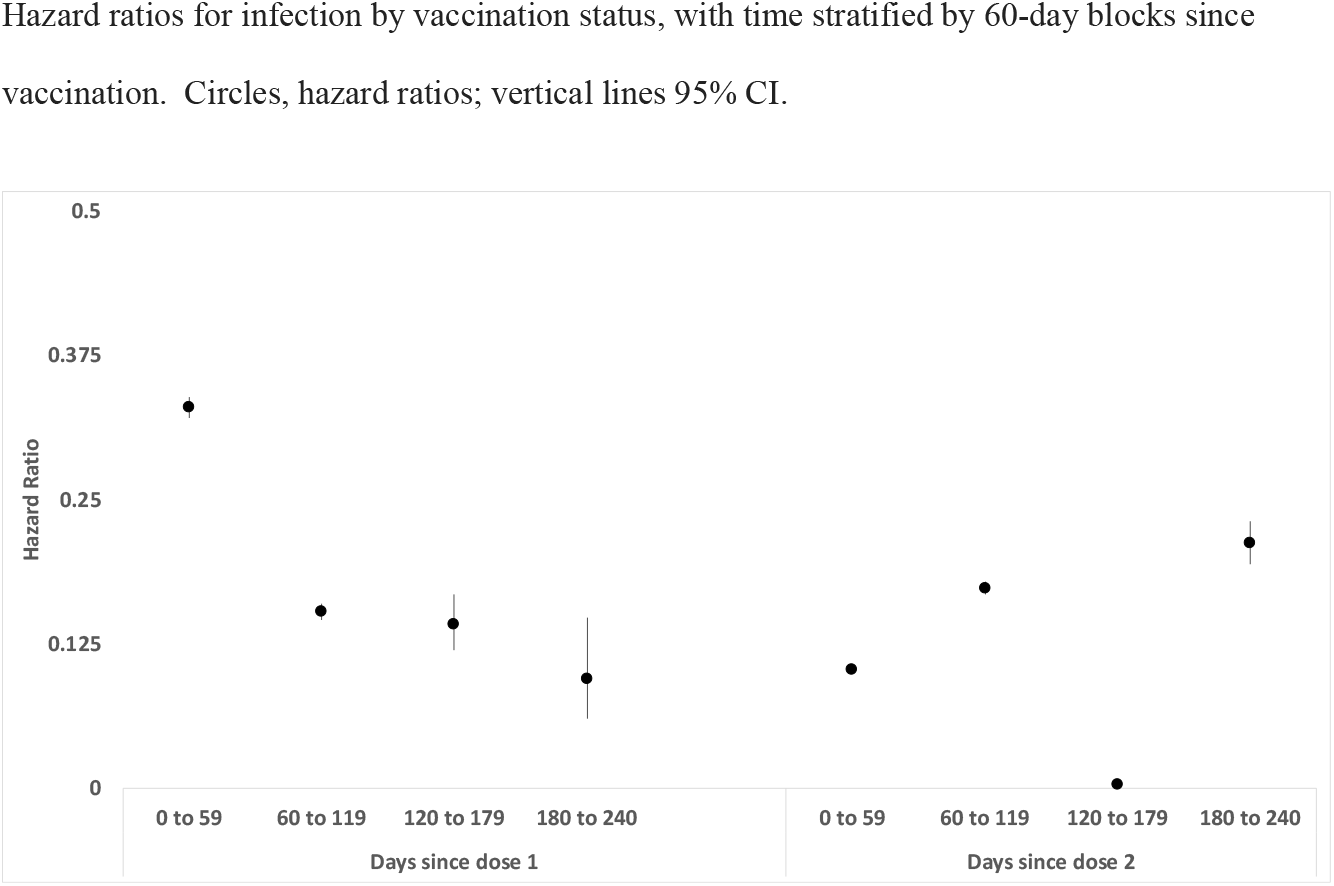
Relative Hazard of Infection by Time Since First or Second SARS-CoV-2 Vaccine Dose. Hazard ratios for infection by vaccination status, with time stratified by 60-day blocks since vaccination. Circles, hazard ratios; vertical lines 95% CI.

## Discussion

We find that in a large cohort of vaccinated individuals in Ontario, Canada vaccination provided remarkable protection against breakthrough infection, with increased protection afforded by a second vaccine dose relative to a single dose. We also find that maximal protection against infection is provided 4-6 months after the second dose of vaccine, at which time protection against infection was nearly complete. However, we found that infection risk increased again after 6 months, suggesting the need for third vaccine doses at this interval (11). Consistent with effects demonstrated using viral neutralization assays (10), we find that the extended interval between first and second vaccine doses has resulted in enhanced immune protection, with a relative reduction in risk of approximately 40% when second vaccine doses were provided 6-8 weeks after first doses. We also identified significant protection against infection associated with heterologous vaccine schedules, in particular the use of initial dose ChAdOx1-S, followed by a second dose of mRNA vaccine, though viral vector vaccines are no longer in wide use in Canada due to concerns about risk of vaccine induced thrombotic thrombocytopenia (VITT)(7). However, the observed effects have substantial face validity and are consistent with the previously described immune enhancement seen with heterologous vaccination schedules as well as the high efficacy of mRNA booster vaccination following non-mRNA SARS-CoV-2 vaccination (27-31).

Our findings on vaccine efficacy are consistent with those reported by others using more typical cohort designs for study of vaccine efficacy. Our application of a case-cohort design, with both cases and controls contributing person-time to unvaccinated, and single- and double-dose vaccinated exposures, allowed us to generate these estimates without access to an unvaccinated control group. The fact that all individuals in our study chose to be vaccinated potentially eliminates confounding by factors associated with vaccination choices.

Like any observational study, ours is potentially subject to confounding by measured and unmeasured factors, though inclusion of available covariates in multivariable models did not significantly change our estimates of vaccine effectiveness. We were also surprised at the sparse nature of data on healthcare worker status, congregate living status and priority health conditions in the CoVaxON dataset. The small number of individuals with these classifications suggests that the dataset is insensitive for these elements. However, inasmuch as CoVaxON data would have been recorded without knowledge of a case’s subsequent infection status these missing data should not result in bias.

In summary, we use a case-cohort study with stratified person-time analysis of vaccination status to demonstrate the strong protection afforded by SARS-CoV-2 vaccines in Ontario, Canada. Despite this strong protection, we do find that an increase in hazard greater than 6 months after second dose of vaccine; our analysis also suggests that increased intervals between vaccine doses, as widely implemented in Canada, provides more durable protection against infection than shorter dosing intervals.

## Data Availability

Data are property of the Government of Ontario. Questions about data and analysis should be directed to the study authors.

## Acknowledgements

The authors with to thank the staff at Public Health Ontario and Ontario’s public health units for collecting, sequencing, analyzing, and providing access to the data used for this analysis.

---

**Supplementary Table.** Multivariable Cox Proportional Hazards Model with Stratified Time Since Vaccination.

| Covariate | Hazard Ratio | 95% Confidence Interval |  |  | P-value |
| --- | --- | --- | --- | --- | --- |
| Time since vaccination |  |  |  |  |  |
| Dose 1 |  |  |  |  |  |
| 0 to 59 | 0.330 | 0.320 | to | 0.339 | <0.001 |
| 60 to 119 | 0.152 | 0.145 | to | 0.159 | <0.001 |
| 120 to 179 | 0.141 | 0.120 | to | 0.167 | <0.001 |
| 180 to 240 | 0.094 | 0.060 | to | 0.148 | <0.001 |
| Dose 2 |  |  |  |  |  |
| 0 to 59 | 0.103 | 0.100 | to | 0.106 | <0.001 |
| 60 to 119 | 0.173 | 0.167 | to | 0.178 | <0.001 |
| 120 to 179 | 0.003 | 0.001 | to | 0.005 | <0.001 |
| 180 to 240 | 0.212 | 0.194 | to | 0.231 | <0.001 |
| Age group |  |  |  |  |  |
| 0-19 years | 0.590 | 0.565 | to | 0.616 | <0.001 |
| 20-49 years |  | 1 (referent) |  |  |  |
| 50-64 years | 0.925 | 0.904 | to | 0.947 | <0.001 |
| 65+ years | 0.753 | 0.732 | to | 0.774 | <0.001 |
| Male sex | 0.995 | 0.977 | to | 1.014 | 0.595 |
| Congregate living | 3.961 | 3.767 | to | 4.165 | <0.001 |
| Healthcare worker | 1.917 | 1.860 | to | 1.976 | <0.001 |
| Priority health condition | 1.190 | 1.124 | to | 1.260 | <0.001 |
| Dosing interval (weeks) |  |  |  |  |  |
| 0-13 days | 0.721 | 0.170 | to | 3.053 | 0.657 |
| 14-27 days |  | 1 (referent) |  |  |  |
| 28-41 days | 0.740 | 0.707 | to | 0.775 | <0.001 |
| 42-55 days | 0.672 | 0.641 | to | 0.705 | <0.001 |
| 56-69 days | 0.825 | 0.789 | to | 0.863 | <0.001 |
| 70-83 days | 0.954 | 0.911 | to | 0.998 | 0.042 |
| 84 days or more | 1.302 | 1.247 | to | 1.359 | <0.001 |
| Vaccine combination (%) |  |  |  |  |  |
| mRNA-mRNA |  | 1 (referent) |  |  |  |
| AZ-AZ | 1.389 | 1.317 | to | 1.466 | <0.001 |
| AZ-mRNA | 0.900 | 0.864 | to | 0.939 | <0.001 |
| Other | 1.578 | 1.503 | to | 1.657 | <0.001 |

**NOTE:** mRNA, Pfizer/BNT or Moderna mRNA vaccine; AZ, Oxford AstraZeneca or Covidshield vaccine (ChAdOx1-S); 95% CI, confidence interval

